# Longitudinal Pain Profiles and Their Clinical and Genetic Correlates in 624,996 U.S. Veterans

**DOI:** 10.64898/2026.10.10.26365209

**Authors:** Christal N. Davis, Mingjian Shi, Tommy Gunawan, Sylvanus Toikumo, Million Veteran Program, Joshua C. Gray, Henry R. Kranzler

**Affiliations:** Mental Illness Research, Education and Clinical Center, Crescenz VAMC, Philadelphia, PA 19104, USA; Center for Studies of Addiction, University of Pennsylvania Perelman School of Medicine, Philadelphia, PA 19104, USA; Department of Medical and Clinical Psychology, Uniformed Services University, Bethesda, MD, 20814, USA; Henry M. Jackson Foundation for the Advancement of Military Medicine Inc., Bethesda, MD, 20817, USA; Human Psychopharmacology Laboratory, National Institute on Alcohol Abuse and Alcoholism, National Institutes of Health, Bethesda, MD, 20892, USA; Office of the Clinical Director, National Institute on Alcohol Abuse and Alcoholism, National Institutes of Health, Bethesda, MD-20892, USA; Regeneron Genetics Center, Tarrytown, NY, 10591, USA

**Author notes:** **Corresponding authors:** Christal N. Davis, Ph.D. and Henry R. Kranzler, M.D., 3535 Market Street, Philadelphia, PA 19104.

**Keywords:** chronic pain, pain profiles, latent profile analysis, electronic health records, polygenic scores, depression, insomnia, Million Veteran Program

## Abstract

**Importance:** Pain varies in its intensity and course, yet much of our understanding of pain is based on cross-sectional assessments or longitudinal studies of selected clinical populations. Routinely collected electronic health record data may enable characterization of longitudinal pain patterns at scale.

**Objective:** To identify longitudinal pain profiles among U.S. Veterans and characterize their clinical and polygenic correlates.

**Design:** Cohort study using electronic health record and genomic data from the Million Veteran Program, with up to 16 years of pain assessments collected between 2000 to 2025.

**Setting:** U.S. Department of Veterans Affairs health care system.

**Participants:** Million Veteran Program participants assigned to African-like, Admixed American-like, East Asian-like, European-like, or South Asian-like genetically inferred ancestry groups who have at least three years of outpatient numeric rating scale pain records.

**Exposures:** Outpatient pain ratings and polygenic scores for psychiatric, sleep, and substance use traits.

**Main Outcomes and Measures:** Pain profiles were identified using Gaussian finite mixture modeling of pain intensity, directional change, and within-person variability. Profiles were characterized by pain, psychiatric and substance use diagnoses, opioid exposure, body mass index, and polygenic scores.

**Results:** Among 624,996 participants (Mean [SD] age at first pain rating=53.4 [13.5] years; 568,833 [91.0%] male), 51,731,567 outpatient pain ratings were analyzed. Three profiles were identified: stable no pain, stable mild pain, and mild-to-moderate increasing pain. Compared with the stable no pain profile, the increasing pain profile had higher odds of chronic pain (OR=21.14 [95% CI: 18.96-23.57]), opioid use disorder (OR=9.32 [95% CI: 8.31-10.45]), opioid exposure (OR=8.61 [95% CI: 8.41-8.83]), major depressive disorder (OR=3.77 [95% CI: 3.67-3.87]), and posttraumatic stress disorder (OR=3.72 [95% CI: 3.61-3.84]). Associations were attenuated but remained after adjustment for mean pain severity. Polygenic scores for depression, insomnia, attention-deficit/hyperactivity disorder, and tobacco use disorder were the most consistent predictors of increasing pain across the three largest genetically inferred ancestry groups.

**Conclusions and Relevance:** Longitudinal modeling of routinely collected pain scores identified pain profiles associated with clinical burden and polygenic liability. These profiles captured clinically meaningful information beyond mean pain severity alone, highlighting the value of longitudinal pain phenotyping in electronic health record data.

**Key Points:** *Question:* Can pain scores collected in outpatient care identify clinically meaningful pain profiles among U.S. Veterans?

*Findings:* In this cohort study of 624,996 Veterans with more than 51 million pain ratings across up to 16 years, three pain profiles were identified: stable no pain, stable mild pain, and mild-to-moderate increasing pain. Profiles differed in clinical burden even after accounting for mean pain severity and were associated with polygenic liability for psychiatric, sleep, and substance use traits.

*Meaning:* Routinely collected pain scores can be used to characterize heterogeneity in the course of pain and factors associated with more burdensome pain patterns.

## Introduction

In 2023, 24.3% of U.S. adults reported chronic pain, and 8.5% reported pain that frequently limited life or work activities^1^. Pain-related healthcare expenditures and lost productivity are estimated to cost hundreds of billions of dollars annually in the U.S.^2,3^. Yet pain is not a static or uniform experience. It may remain mild, persist at high severity levels, progressively worsen, or fluctuate over time^4–8^. Understanding this heterogeneity is important for identifying individual differences in the long-term burden of pain and the factors associated with different clinical courses.

Much of what is known about pain at the population level is based on cross-sectional assessments of pain-related interference, chronic pain status, or summary measures of pain severity^9^. Although highly informative, these approaches obscure meaningful variation over time, and individuals with similar average pain levels may have markedly different longitudinal experiences. Characterizing these patterns and identifying factors that distinguish them could improve our ability to identify individuals at risk for persistent or worsening pain and inform earlier, more targeted interventions.

Longitudinal studies have demonstrated that the course of pain is heterogeneous. Across clinical pain populations, investigators have identified stable low-pain, persistent, improving, worsening, episodic, and fluctuating courses^4–8^. However, most studies focused on specific pain conditions, included relatively modest sample sizes, or followed participants over comparatively short periods^6,7^. Recently, a large international study of over 6 million individuals established population-level reference curves for pain across the lifespan, characterizing variation in pain burden by age, sex, and geographic region^10^. However, important questions remain about how pain evolves within individuals over time, including what distinct longitudinal courses emerge and how these relate to differences in clinical burden and genetic liability. Routinely collected electronic health record (EHR) data offer an opportunity to address this gap by leveraging repeated pain assessments collected over many years.

Longitudinal phenotyping in EHR data also enables the identification of clinical and sociodemographic characteristics that differentiate the course of pain and are associated with more burdensome pain patterns. Pain commonly co-occurs with depression, anxiety, and sleep disturbances^11,12^, and understanding how these factors differ across longitudinal pain profiles may provide insight into the clinical context of persistent or worsening pain^13^. Genetic liability may also distinguish pain profiles. Although genomic studies indicate shared liability among chronic pain, major depression, and insomnia^14–16^, whether these liabilities are associated with the long-term course of pain is unknown.

We used longitudinal EHR data from the Million Veteran Program (MVP) to identify pain profiles over a 16-year follow-up. To test whether the pain profiles capture clinically meaningful differences, we assessed the prevalence of pain diagnoses, psychiatric diagnoses, opioid exposure, and body mass index (BMI) across profiles. We then tested whether psychiatric, sleep, and substance-use polygenic scores (PGS) predicted pain profile membership across genetically inferred ancestry (GIA) groups.

## Methods

### Study Design, Participants, and Ethics

The MVP is a national biobank implemented by the U.S. Department of Veterans Affairs^17^. The MVP was approved by the VA Central Institutional Review Board (IRB) and local site IRBs. Participants provide a blood sample for genomic analysis and informed consent to link their genomic data with their VA EHR data. Genotyping and quality-control procedures have been described previously^18^.

Initial data comprised 110,479,848 numeric rating scale pain scores from 641,630 participants assigned to African (AFR), Admixed American (AMR), East Asian (EAS), European (EUR), or South Asian (SAS) GIA groups. Ratings were recorded on a 0-10 scale at outpatient and inpatient VA visits. We excluded records from before 2000, values >10 or non-integers, ratings recorded during an inpatient stay or on the admission or discharge date to minimize the capture of acute pain incidents, ratings obtained before age 18 or after age 100, and participants with fewer than three years of pain records (see Figure 1). The final dataset comprised 51,731,567 outpatient pain ratings from 624,996 participants. Because participant density declined in the later follow-up years (Supplementary Figure 1), data were capped at 16 years from the first assessment based on the median record length for the sample. These procedures yielded 7,774,115 person-years of data.

**Figure 1.**
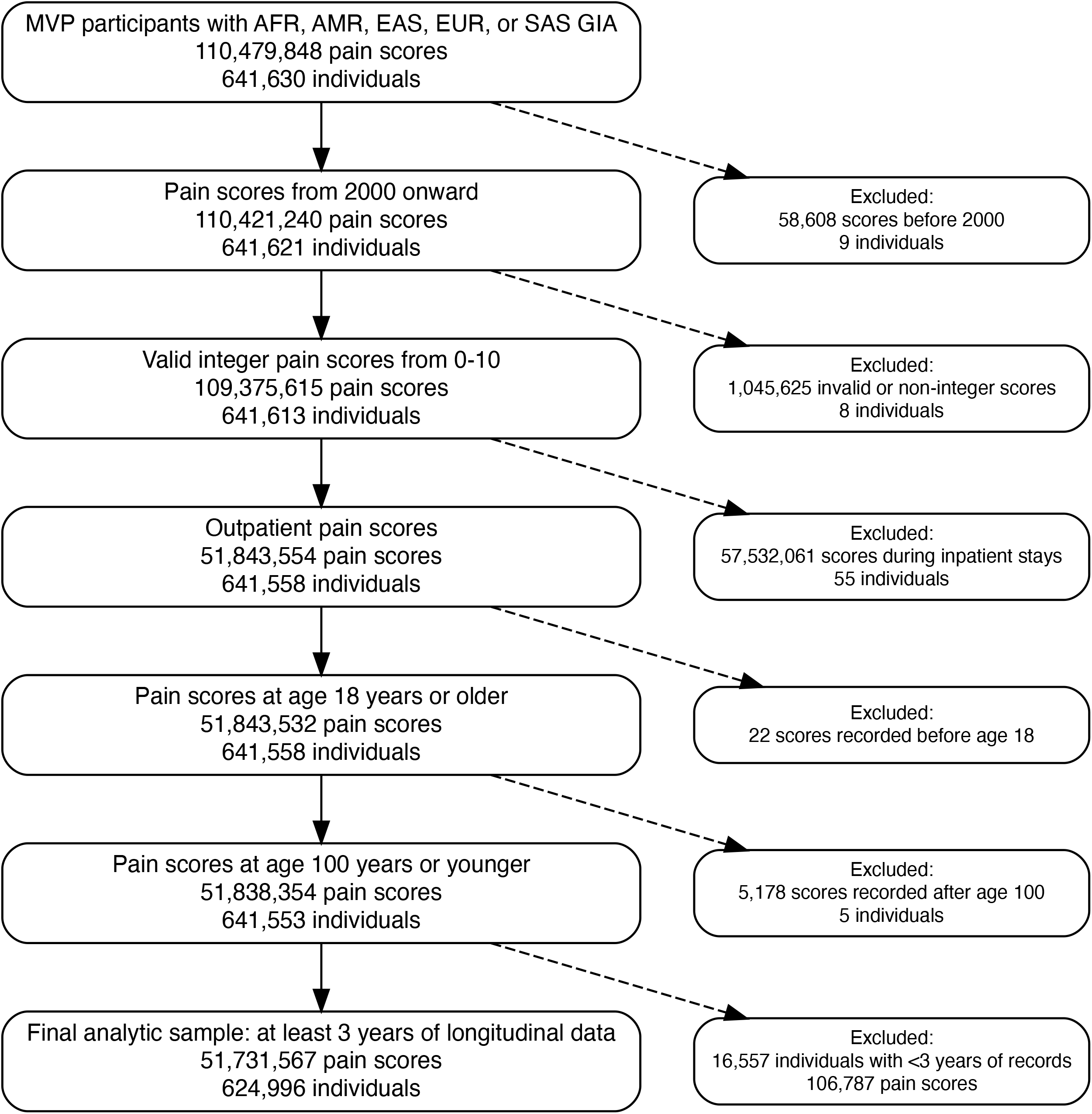
Sample inclusion and exclusion criteria. *Note:* MVP = Million Veteran Program, AFR = African, AMR = Admixed American, EAS = East Asian, EUR = European, SAS = South Asian, and GIA = genetically inferred ancestry.

### Latent Profile Modeling

Using person-years data, we calculated three measures of annual pain intensity: mean, median, and maximum pain severity. Because these measures were highly correlated and including all three could overweight pain intensity when deriving profiles, we used principal component analysis to obtain a composite measure of pain intensity. The first principal component explained 88.45% of the variance across the three measures (loadings of 0.61 for mean pain, 0.58 for median pain, and 0.54 for maximum pain) and was retained. We also calculated (1) change in pain as the linear slope from an ordinary least squares regression of annual mean pain on time in years since the first assessment and (2) within-person pain instability as the root mean square of successive differences (RMSSD) across ordered annual mean pain scores. These three features, which represent pain intensity, directional change, and temporal instability, were included in profile modeling.

Gaussian finite mixture models were fitted to the three features using model-based clustering^19^. Solutions specifying 3 through 8 profiles were compared using the Bayesian Information Criterion, classification entropy, average posterior probability, profile size, and clinical interpretability (Supplementary Table 1). The selected three-profile solution used a VVE (variable volume, variable orientation, equal shape) covariance structure. The solution had a mean maximum posterior probability of 0.94 and normalized entropy of 0.84. Profiles were labeled as “stable no pain,” “stable mild pain,” and “mild-to-moderate increasing pain” based on their longitudinal pain patterns. The stable no pain profile served as the reference group in subsequent primary regression models; secondary models used stable mild pain as the reference to directly compare the two profiles characterized by pain.

### Clinical Characterization of Profiles

We characterized individuals within each pain profile by examining their demographic and clinical characteristics, including age, sex, number of years of data, GIA assignment, body mass index (BMI), mean Alcohol Use Disorders Identification Test-Consumption (AUDIT-C) score, pain-related diagnoses, psychiatric diagnoses, substance use disorders (SUDs), and opioid exposure. Pain-related diagnoses included back pain, widespread pain, chronic pain, pelvic pain, neck/shoulder pain, limb and joint pain, arthritis and arthrosis, osteoarthritis, diabetic neuropathy, fibromyalgia, complex regional pain syndrome, and other pain-related conditions. Psychiatric and SUDs included major depressive disorder, posttraumatic stress disorder, anxiety disorders, insomnia, alcohol use disorder, tobacco use disorder, cannabis use disorder, and opioid use disorder. The ICD-9 and ICD-10 codes for these diagnoses are provided in Supplementary Table 2.

We first examined differences across profiles using unadjusted chi-square tests and analysis of variance. We then conducted regression models (logistic for binary variables and linear for continuous variables) to estimate associations between profile membership and clinical and demographic characteristics after adjusting for age, sex, GIA assignment, and number of years of pain data. Finally, we repeated the regression models including participant-level mean pain as an additional covariate to determine whether profile membership remained independently associated with clinical characteristics.

### Polygenic Score Associations

We evaluated 13 PGS indexed in the PGS Catalog^20^: feelings of worry or anxiety (PGS001021), other anxiety disorders (PGS004451), depression (PGS004759), alcohol intake frequency (PGS001087), drinks per week from EUR and AFR discovery cohorts (PGS003376 and PGS003373), ever having used cannabis (PGS002125), schizophrenia (PGS000133), attention-deficit/hyperactivity disorder (ADHD; PGS002746), bipolar I disorder (PGS002787), bipolar II disorder (PGS002788), sleeplessness/insomnia (PGS002149), and tobacco use disorder (PGS004450). PGS were standardized within each GIA group before analysis.

Association analyses were conducted separately within each GIA group. Each PGS was entered into a separate multinomial logistic regression model predicting profile membership, adjusting for age, sex, number of years of pain data, and the first 10 within-ancestry genetic principal components. The Benjamini-Hochberg procedure was used to control the false discovery rate (FDR) across the 26 PGS tests within each GIA group, and adjusted p-values or q values are reported.

## Results

### Cohort Characteristics

As shown in Table 1, the mean age at first pain rating was 53.41 years (SD, 13.53), and 568,833 participants (91.02%) were male. GIA assignments were EUR (n=437,583; 70.01%), AFR (n=121,030; 19.36%), AMR (n=59,093; 9.45%), EAS (n=6,734; 1.08%), and SAS (n=556; 0.09%). The mean pain score was 2.26 (SD=1.70), with a median of 61 ratings per participant (mean=82.77; SD=115.82). The mean uncapped record span was 15.92 years (SD=5.63; median=16 years). Mean documented pain increased gradually over calendar time, was consistently higher among female than male participants, and differed across GIA groups (Supplementary Figures 2-4).

**Table 1.** Sample characteristics.

| **Characteristic** | **Mean / Median (SD) or % (N)** |
| --- | --- |
| Age at first pain score | 53.41 / 55 (13.53) |
| Sex* |  |
| Male | 91.02% (568,833) |
| Female | 8.98% (56,147) |
| Genetically inferred ancestry |  |
| AFR | 19.36% (121,030) |
| AMR | 9.45% (59,093) |
| EAS | 1.08% (6,734) |
| EUR | 70.01% (437,583) |
| SAS | 0.09% (556) |
| Person-level average pain score | 2.26 / 1.89 (1.70) |
| Person-level average pain severity |  |
| No pain | 2.58% (16,138) |
| Mild (>0 to <4) | 79.83% (498,949) |
| Moderate (4 to <7) | 16.90% (105,642) |
| Severe (≥7) | 0.68% (4,267) |
| Number of pain scores per person | 82.77 / 61 (115.82) |
| Uncapped length of record (years) | 15.92 / 16 (5.63) |
| *Note: AFR = African-like, AMR = Admixed American-like, EAS = East Asian-like, EUR = European-like, SAS = South Asian-like.* |  |
| *\* 16 individuals were missing information on sex.* |  |

### Longitudinal Pain Profiles

The three-profile solution identified stable no pain, stable mild pain, and mild-to-moderate increasing pain profiles (Figure 2). Pain ratings in the stable no pain group remained near zero throughout follow-up (M=0.07), with 87.9% of annual pain scores equal to zero and little within-person variability (RMSSD=0.21). The stable mild pain group showed low-level pain (M=0.87) with little directional change. Approximately half of annual pain scores (50.2%) were zero, and there was moderate within-person variability (RMSSD=1.48). In contrast, the mild-to-moderate increasing profile began with a mean yearly pain score of ~3 and rose steadily across follow-up (M=3.27, maximum=6.80), showing the greatest (although still modest) increase in pain over time (mean slope=0.056 points/year), and within-person variability (RMSSD=2.55). Only 18.6% of annual pain scores in this profile were zero.

### Profile Characterization

Clinical and demographic characteristics differed across the three pain profiles in unadjusted analyses (all *p*<.001), with the mild-to-moderate increasing pain profile showing the greatest burden of pain-related diagnoses, psychiatric disorders, SUDs, opioid exposure, and higher BMI (Supplementary Tables 3-6). In regression models adjusted for age, sex, GIA, and number of years of pain data, membership in the stable mild and mild-to-moderate increasing pain profiles was associated with higher odds of all examined clinical conditions relative to the stable no pain profile (Figure 3; Supplementary Tables 7-8). Additional adjustment for participant-level mean pain attenuated these effects, but both pain profiles remained associated with higher odds of all examined clinical conditions relative to the stable no pain profile (Figure 3; Supplementary Tables 9-10). Associations with BMI also persisted after adjustment for mean pain, whereas the association between stable mild pain and mean AUDIT-C was no longer significant. In secondary analyses with stable mild pain as the reference group (Supplementary Tables 11-14), the increasing profile remained associated with greater clinical burden even after adjustment for mean pain, including having higher odds of chronic pain (OR=1.65; 95% CI: [1.60-1.70]), opioid use disorder (OR=1.48; 95% CI: [1.42-1.54]), and PTSD (OR=1.22; 95% CI: [1.20-1.24]).

**Figure 2.**
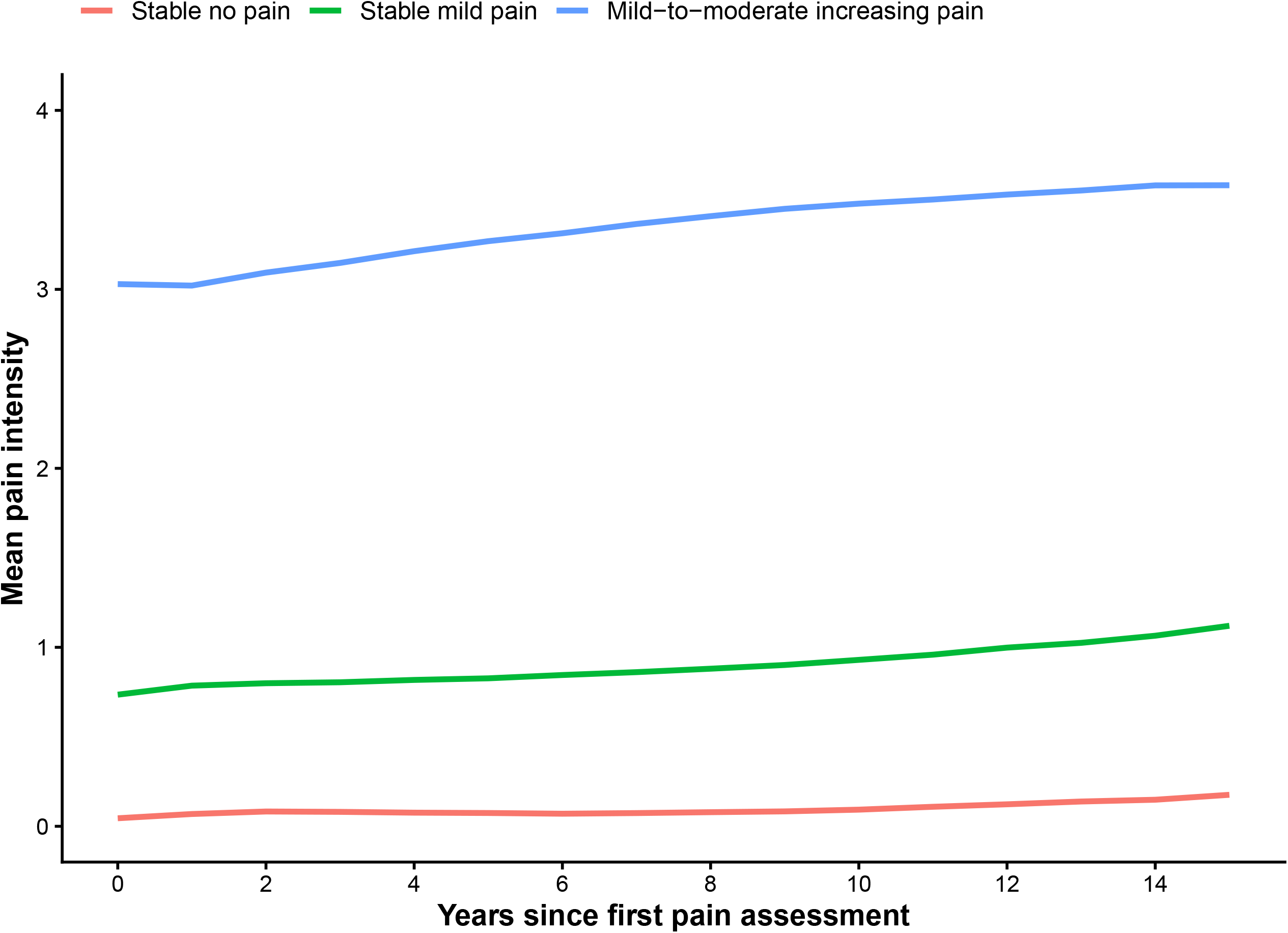
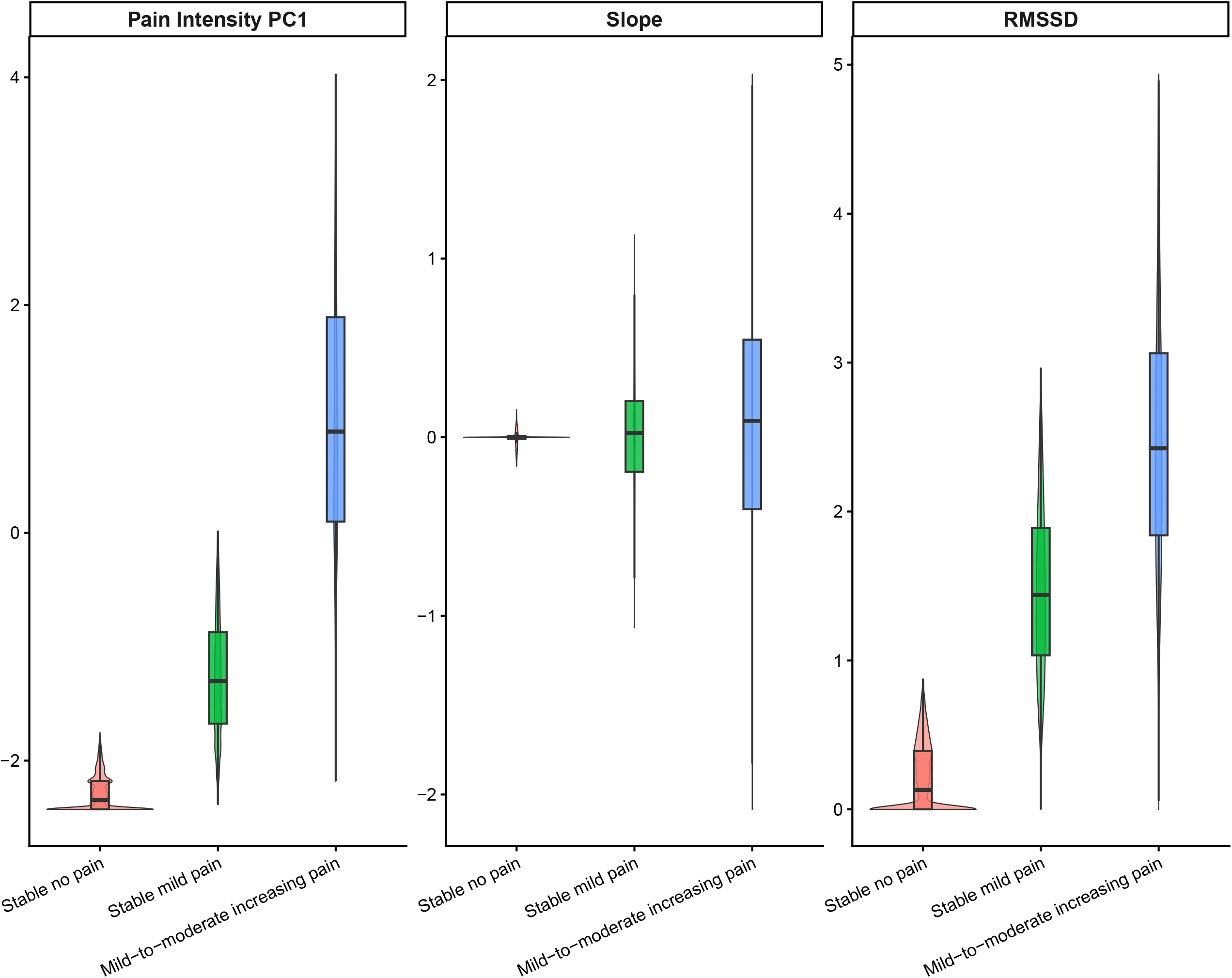
Longitudinal pain profiles. *Note:* Panel A shows the mean severity of pain across profiles over the follow-up period. Panel B shows the distribution of model features for each longitudinal pain profile. PC1 = first principal component of a pain intensity composite comprised of mean, median, and maximum annual pain, RMSSD = root mean square of successive differences. Values on the features in Panel B are capped at the 1^st^ and 99^th^ percentile for visualization.

**Figure 3.**
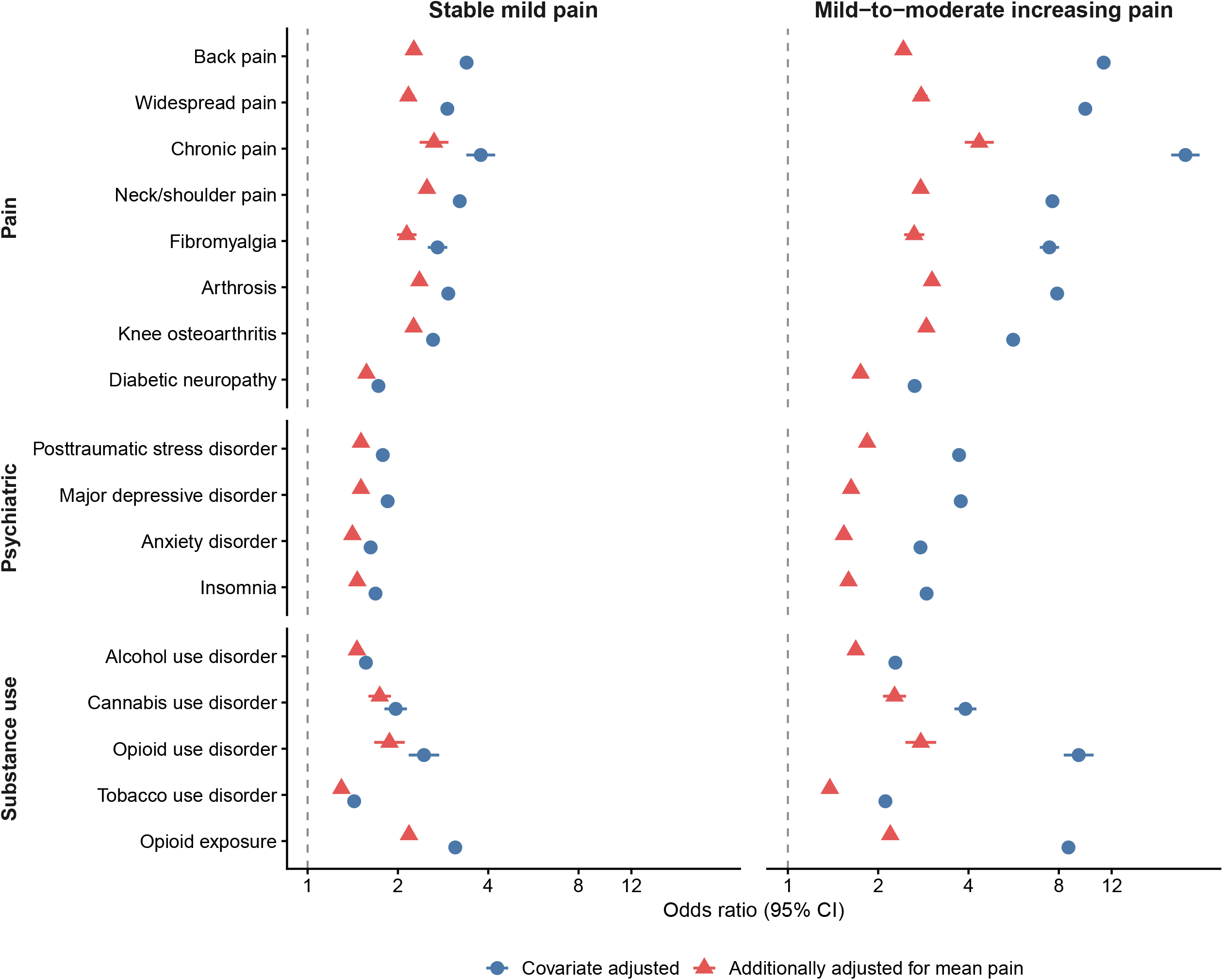
Clinical characterization of pain profiles. *Note:* Odds ratios and 95% confidence intervals are shown for selected pain, psychiatric, and substance use outcomes comparing the stable mild pain and mild-to-moderate increasing pain profiles with the stable no pain profile. Covariate-adjusted models included age, sex, genetically inferred ancestry, and number of years of available pain data, while mean-pain-adjusted models additionally included participant-level mean pain as a covariate. Complete results for all clinical characteristics are provided in Supplementary Tables 7-10.

### PGS Associations

Depression PGS showed the most consistent positive associations with pain profile membership across GIA groups (Figure 4; Supplementary Table 15). In EUR participants, higher depression PGS was associated with both stable mild pain (β=0.116; q=9.71×10^-84^) and mild-to-moderate increasing pain (β=0.233; q<1×10^-300^) relative to stable no pain. Similar associations were observed in AFR (stable mild: β=0.048; q=.016; increasing: β=0.094; q=4.24×10^-8^) and AMR participants (stable mild: β=0.098; q=4.73×10^-5^; increasing: β=0.186; q=1.50×10^-16^). Depression PGS was not a significant predictor in EAS participants, whereas in SAS participants it was associated only with increasing pain (β=0.940; q=.005). In secondary analyses using stable mild pain as the reference group (Supplementary Table 16), higher depression PGS remained associated with increasing pain in EUR (β=0.117; q=2.40×10^-275^), AFR (β=0.046; q=1.07×10^-10^), and AMR participants (β=0.087; q=3.40×10^-19^).

**Figure 4.**
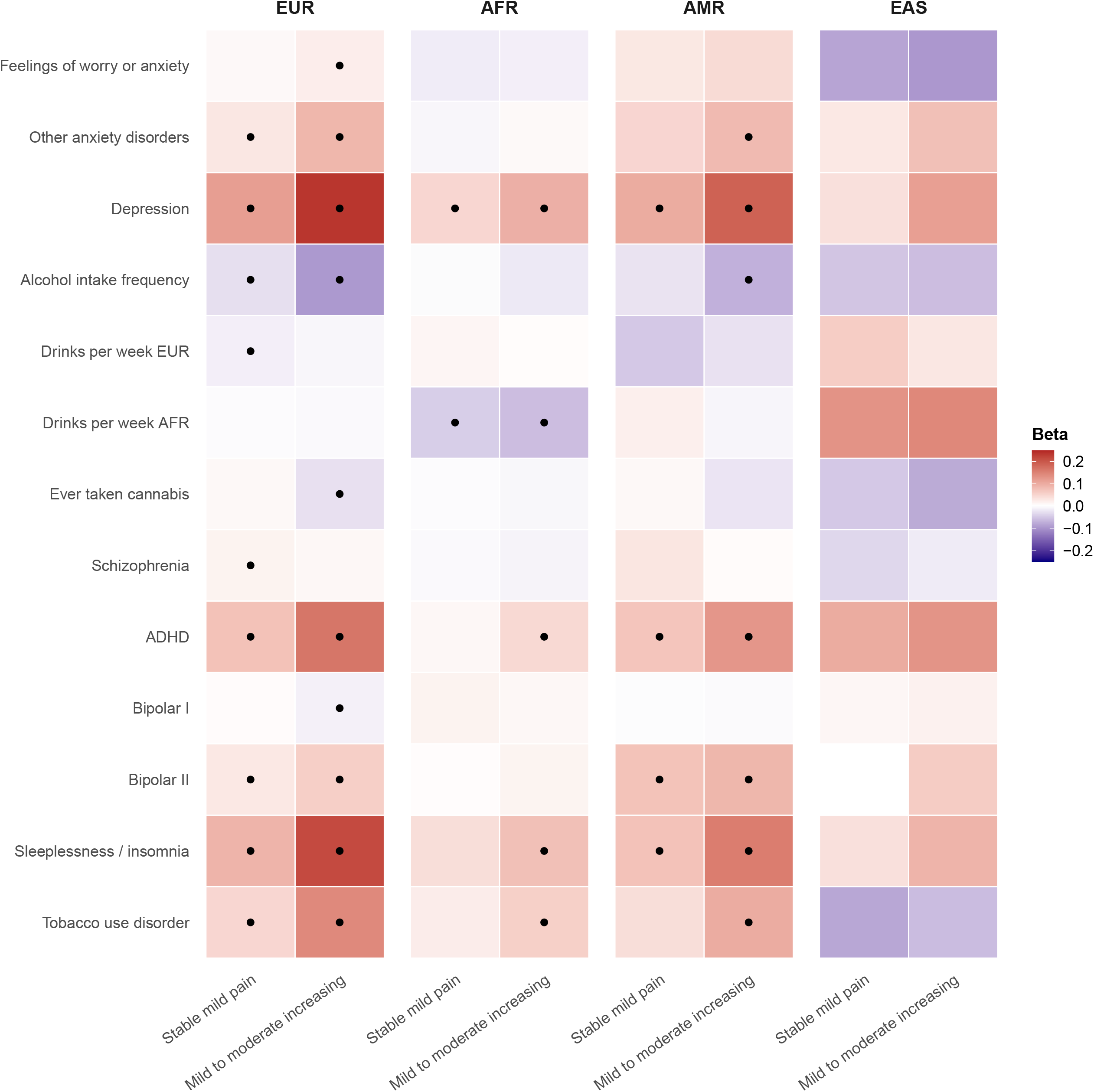
Polygenic score associations with pain profile membership across genetically inferred ancestry groups. *Note:* The South Asian (SAS) genetically inferred ancestry group is not shown due to imprecise estimates resulting from the smaller sample size. In SAS, the only significant association after false discovery rate (FDR) correction was between higher depression polygenic scores (PGS) and greater likelihood of membership in the increasing pain profile relative to the stable no pain profile. PGS are presented on the Y-axis, and the stable no pain profile was the reference group in all multinomial models. Dots represent associations that were significant after FDR correction. EUR = European, AFR = African, AMR = Admixed American, and EAS = East Asian, ADHD = attention deficit hyperactivity disorder. Genetically inferred ancestry groups are presented in order of their prevalence in the Million Veteran Program.

Several other PGS were associated with pain profiles, particularly in EUR participants (Figure 4). Sleeplessness/insomnia, ADHD, and tobacco use disorder PGS were positively associated with both profiles relative to stable no pain in EUR. In AMR participants, sleeplessness/insomnia and ADHD PGS were similarly associated with both profiles, whereas tobacco use disorder PGS was associated only with increasing pain. In AFR participants, sleeplessness/insomnia, ADHD, and tobacco use disorder PGS were associated with increasing pain but not stable mild pain. In secondary analyses, these three PGS also distinguished increasing from stable mild pain in EUR (β=0.123, 0.091, and 0.092, respectively; all q<1×10^-167^), AFR (β=0.036, 0.035, and 0.034; all q<2×10^-6^), and AMR participants (β=0.082, 0.055, and 0.058; all q<5×10^-8^). No PGS associations survived FDR correction in EAS participants.

Alcohol-related PGS showed heterogeneous associations (Figure 4). Alcohol intake frequency PGS was inversely associated with both pain profiles relative to stable no pain in EUR participants and with increasing pain in AMR participants. The AFR-derived drinks per week PGS was inversely associated with both pain profiles relative to stable no pain in AFR participants. Associations for the drinks-per-week PGS were ancestry-specific. In secondary analyses (Supplementary Table 16), alcohol intake frequency PGS was inversely associated with increasing versus stable mild pain in EUR (β=−0.067; q=1.08×10^-90^), AFR (β=−0.017; q=.033), and AMR participants (β=−0.048; q=1.93×10^-6^). Other PGS associations varied across profiles and GIA group.

## Discussion

Among 624,996 U.S. Veterans with more than 51 million pain ratings across up to 16 years, we identified three profiles characterized by stable no pain, stable mild pain, and mild-to-moderate increasing pain. Profiles differed substantially in the clinical burden of pain and co-occurring medical, psychiatric, and substance use disorders, and these associations persisted after accounting for pain severity, indicating that longitudinal pain course captures clinically meaningful information beyond average pain severity. PGS for psychiatric, sleep, and substance use traits also differentiated pain profiles, including increasing versus stable mild pain. Together, these findings demonstrate the value of characterizing the longitudinal course of pain at scale and suggest that pain patterns reflect differences in clinical and genetic liability.

Stable, persistent, recovering, worsening, and fluctuating patterns have been identified in clinical pain populations^2,3,8,17,21^. Here, we extended this work to a large, clinically heterogeneous population with pain ratings collected during routine outpatient care. Across the three profiles we identified, there was a clear gradient in the associated clinical burden. Relative to participants in the stable no pain profile, those in the stable mild pain profile and, particularly, the mild-to-moderate increasing pain profile had more pain diagnoses, psychiatric and substance use disorders, prescription opioid exposure, and higher BMI. Previous longitudinal pain studies also identified poorer mental health and greater multimorbidity among individuals with more burdensome pain profiles^6,22,23^. Although adjusting for mean pain severity attenuated the associations, the increasing profile remained associated with greater clinical burden even when directly compared with stable mild pain. These findings underscore how individuals with similar pain levels may experience substantially different longitudinal courses and clinical burdens.

PGS associations demonstrated that the observed profiles are related to genetic liability for psychiatric and sleep conditions. PGS for depression, sleeplessness/insomnia, ADHD, and tobacco use disorder were the most consistent predictors of the mild-to-moderate increasing pain profile across the three largest GIA groups. The depression and insomnia findings are consistent with the documented comorbidity between these conditions and chronic pain^15,24^, and with genomic evidence for their shared liability^24,25^. Similarly, previous studies have identified substantial genomic overlap between ADHD and multisite chronic pain^14^ and between chronic pain and SUDs^15^. Importantly, depression, sleeplessness/insomnia, ADHD, and tobacco use disorder PGS also distinguished mild-to-moderate increasing from stable mild pain across EUR, AFR, and AMR participants. These findings suggest that genetic liability for these traits is associated not only with the presence of pain but also with differences in longitudinal pain course among individuals experiencing pain.

The alcohol-related PGS findings were more heterogeneous. In primary analyses, PGS for higher alcohol intake frequency were associated with lower odds of belonging to the stable mild and increasing pain profiles than the stable no pain profile in EUR participants, and in AFR individuals higher PGS for drinks per week showed similar associations. Alcohol intake frequency PGS also consistently distinguished increasing from stable mild pain in EUR, AFR, and AMR participants, with higher PGS being associated with lower likelihood of the increasing profile. Although these associations may appear counterintuitive, they align with evidence that the relationship between alcohol and pain differs for consumption versus problematic use and alcohol use disorder. Observational studies have reported inverse associations between alcohol consumption and chronic pain^26^, while AUD is associated with greater pain burden^27^. In recent studies, although a general chronic pain factor was positively genetically correlated with a broad addiction factor, after accounting for shared addiction liability, the pain factor was negatively genetically correlated with alcohol consumption frequency^28^. Importantly, the inverse associations we observed for alcohol consumption PGS should not be interpreted as evidence that alcohol consumption is protective against pain, as genetically informed analyses have not supported a causal protective effect^29^.

These findings should be considered in the context of several limitations. First, pain ratings were collected during routine outpatient care rather than at standardized intervals. This contributes to variation in the timing and frequency of assessments across participants, which may partially reflect differences in an individual’s healthcare utilization and VA care duration. However, annual aggregation of the pain ratings reduced the influence of these differences in assessment frequency. Second, numeric pain ratings capture pain intensity, but not other dimensions of pain, including interference and location. Incorporating these features would provide a more complete characterization of pain over time. Third, the identified profiles depend on the features and modeling approach used, and replication in independent samples and using alternative approaches will be important. Fourth, the clinical association analyses do not establish temporality or causal relationships. Similarly, although PGS measure genetic liability that precedes the observed pain course, they are not informative as to the mechanisms by which that liability is linked with pain. PGS performance also varied across GIA populations because of differences in the availability and size of discovery GWAS and other factors^4,19^. In particular, the smaller EAS and SAS samples limited our ability to evaluate the consistency of associations across GIA groups. Finally, the predominantly older male Veteran sample limits generalizability to female adults, younger populations, and individuals receiving care outside the VA.

## Conclusions

These findings highlight the importance of considering both how much pain an individual experiences and how their pain changes over time. Individuals with similar initial levels of pain may follow substantially different courses, and these differences are associated with overall clinical burden and genetic liability for psychiatric, sleep, and SUDs. Thus, longitudinal pain phenotyping captured meaningful heterogeneity not reflected in average pain severity alone. Leveraging longitudinal information from EHR data may provide a scalable approach to better characterize the course of pain and identify individuals at risk for more burdensome pain and greater morbidity.

## Supporting information

MVP Core Acknowledgment

## Statements

### Data Availability

Access to individual-level Million Veteran Program data is available to VA researchers on approved federally funded projects.

## Acknowledgments

The view(s) expressed herein are those of the author(s) and do not reflect the official policy or position of the Henry M. Jackson Foundation for the Advancement of Military Medicine Inc. The contents of this manuscript also do not represent the views of the U.S. Department of Veterans Affairs, Uniformed Services University, the Department of War, or the U.S. Government.

