## Supplementary Materials for "Longitudinal Pain Profiles and Their Clinical and Genetic Correlates in 624,996 U.S. Veterans"

### **Results of Alternative Reference Group Analyses**

To further characterize differences between the pain profiles, regression models were repeated using the stable mild pain profile as the reference group rather than the stable no pain group. In models adjusted for age, sex, genetically inferred ancestry (GIA) group, and duration of follow-up, Veterans in the mild-to-moderate increasing pain profile had higher odds of all examined pain, medical, psychiatric, and substance use outcomes compared with those in the stable mild pain profile. Associations were largest for chronic pain (OR, 5.60; 95% CI, 5.46-5.73), opioid use disorder (OR, 3.81; 95% CI, 3.70-3.94), widespread pain (OR, 3.35; 95% CI, 3.31-3.40), back pain (OR, 3.33; 95% CI, 3.29-3.37), complex regional pain syndrome (OR, 3.29; 95% CI, 3.06-3.54), fibromyalgia (OR, 2.75; 95% CI, 2.69-2.80), arthrosis (OR, 2.68; 95% CI, 2.65-2.72), and arthritis (OR, 2.56; 95% CI, 2.53-2.59). The mild-to-moderate increasing pain profile also had higher BMI (β, 0.99; 95% CI, 0.95-1.02) and slightly lower mean AUDIT scores (β, −0.09; 95% CI, −0.10 to −0.08) than the stable mild pain profile.

After additional adjustment for mean pain severity, associations were attenuated but remained significant for 31 of 32 binary outcomes. The largest remaining associations with the mild-to-moderate increasing pain profile were observed for chronic pain (OR, 1.65; 95% CI, 1.60-1.70), opioid use disorder (OR, 1.48; 95% CI, 1.42-1.54), cocaine use disorder (OR, 1.37; 95% CI, 1.33-1.42), arthritis (OR, 1.32; 95% CI, 1.30-1.34), carpal tunnel syndrome (OR, 1.31; 95% CI, 1.28-1.34), rheumatoid arthritis (OR, 1.31; 95% CI, 1.25-1.36), and cannabis use disorder (OR, 1.30; 95% CI, 1.26-1.35). Associations also persisted for widespread pain, arthrosis, knee osteoarthritis, hip osteoarthritis, enthesopathy, PTSD, and other examined outcomes. Opioid exposure was the only binary outcome that no longer differed between the mild-to-moderate increasing and stable mild pain profiles after adjustment for mean pain severity (OR, 1.01; 95% CI, 0.99-1.03; P = .44). The mild-to-moderate increasing pain profile continued to have higher BMI (β, 0.35; 95% CI, 0.31-0.40) and had slightly higher mean AUDIT scores (β, 0.03; 95% CI, 0.02-0.04) after adjustment for mean pain severity, although the magnitude was very small.

Compared with the stable mild pain profile, the stable no pain profile had lower odds of all examined binary outcomes in both sets of models. Before adjustment for mean pain severity, odds ratios ranged from 0.26 for chronic pain (95% CI, 0.24-0.30) to 0.70 for nicotine use disorder (95% CI, 0.68-0.72). These differences remained after adjustment for mean pain severity, with odds ratios ranging from 0.38 for chronic pain (95% CI, 0.34-0.42) to 0.77 for nicotine use disorder (95% CI, 0.75-0.79). The stable no pain profile also had lower BMI than the stable mild pain profile before (β, −0.87; 95% CI, −0.93 to −0.81) and after (β, −0.69; 95% CI, −0.75 to −0.63) adjustment for mean pain severity. Mean AUDIT scores were slightly higher in the stable no pain profile before adjustment (β, 0.02; 95% CI, 0.004-0.04) but did not differ after adjustment for mean pain severity (β, −0.01; 95% CI, −0.03 to 0.001; P = .07).

### **Supplementary Figure 1. Density of pain scores across the follow-up period.**

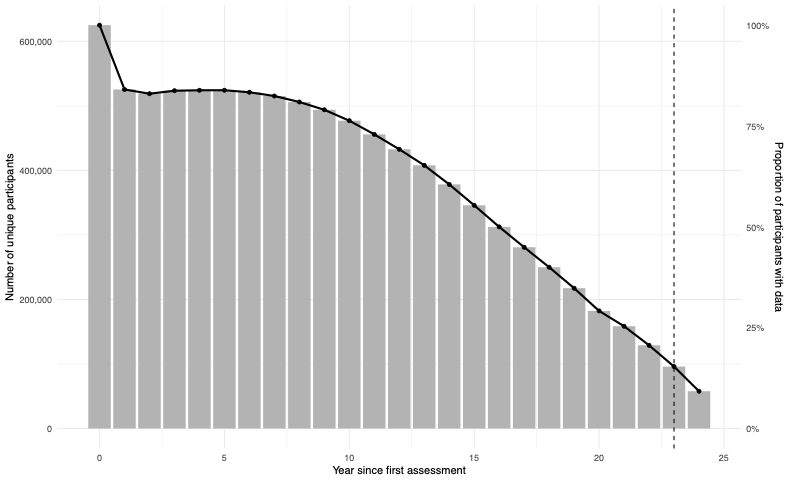

### **Supplementary Figure 2. Mean pain ratings across calendar years.**

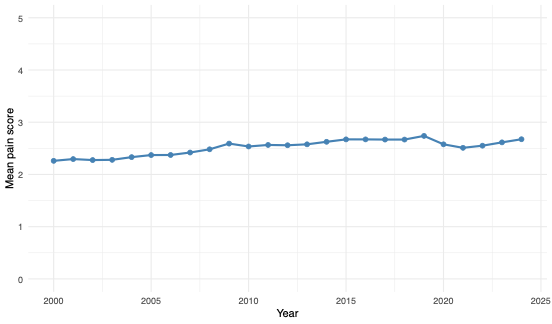

### **Supplementary Figure 3. Mean pain scores across time by sex.**

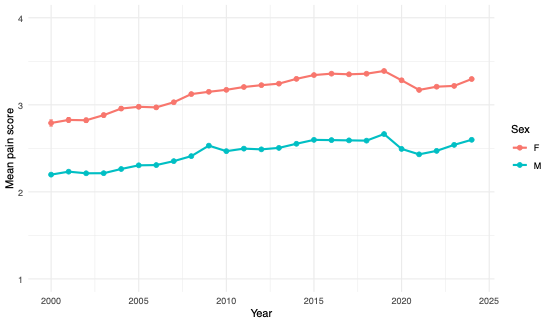

### **Supplementary Figure 4. Mean pain scores across time by genetically inferred ancestry group.**

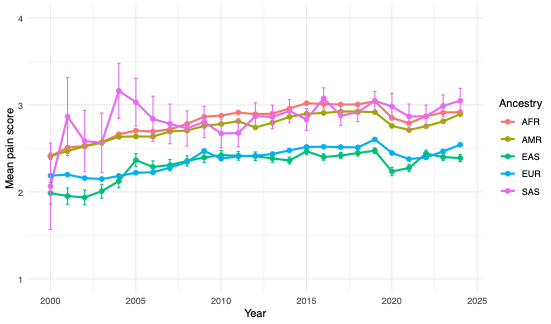
