## Supplementary material for "Longitudinal Pain Profiles and Their Clinical and Genetic Correlates in 624,996 U.S. Veterans": MVP Core Acknowledgment

### VA Million Veteran Program
### Core Acknowledgements for Publications
### June 2026

#### MVP Program Office

- Sumitra Muralidhar, Ph.D., Program Director
  US Department of Veterans Affairs, 810 Vermont Avenue NW, Washington, DC 20420
- Jennifer Moser, Ph.D., Associate Director, Scientific Programs
  US Department of Veterans Affairs, 810 Vermont Avenue NW, Washington, DC 20420
- Jennifer E. Deen, B.S., Associate Director, Cohort & Public Relations
  US Department of Veterans Affairs, 810 Vermont Avenue NW, Washington, DC 20420

#### MVP Steering Committee

- Co-Chair: J. Michael Gaziano, M.D., M.P.H.
  VA Boston Healthcare System, 150 S. Huntington Avenue, Boston, MA 021
- Co-Chair: Dave Oslin, M.D.
  Philadelphia VA Medical Center, 3900 Woodland Avenue, Philadelphia, PA 19104
- Sumitra Muralidhar, Ph.D., Ex-Officio
  US Department of Veterans Affairs, 810 Vermont Avenue NW, Washington, DC 20420
- Drew Helmer, M.D., M.S.
  Michael E. DeBakey VA Medical Center, 2002 Holcombe Boulevard, Houston, TX 77030
- Adriana Hung, M.D., M.P.H.
  VA Tennessee Valley Healthcare System, 1310 24th Avenue, South Nashville, TN 37212
- Philip S. Tsao, Ph.D.
  VA Palo Alto Health Care System, 3801 Miranda Avenue, Palo Alto, CA 94304
- Deepak Voora, M.D.
  Durham VA Medical Center, 508 Fulton Street, Durham, NC 27705

#### MVP Co-Principal Investigators

- J. Michael Gaziano, M.D., M.P.H.
  VA Boston Healthcare System, 150 S. Huntington Avenue, Boston, MA 02130
- Philip S. Tsao, Ph.D.
  VA Palo Alto Health Care System, 3801 Miranda Avenue, Palo Alto, CA 94304

#### MVP Core Operations

- Jessica V. Brewer, M.P.H., Director, MVP Cohort Operations
  VA Boston Healthcare System, 150 S. Huntington Avenue, Boston, MA 02130
- Kelly Cho, M.P.H, Ph.D., Director, MVP Phenomics
  VA Boston Healthcare System, 150 S. Huntington Avenue, Boston, MA 02130
- Lori Churby, B.S., Director, MVP Regulatory Affairs
  VA Palo Alto Health Care System, 3801 Miranda Avenue, Palo Alto, CA 94304
- Yonghui Jia, Ph.D., Director, VA Central Biorepository

VA Boston Healthcare System, 150 S. Huntington Avenue, Boston, MA 02130
- Jacob T. Kean, Ph.D., Acting Director, VA Informatics and Computing Infrastructure (VINCI)
  VA Salt Lake City Health Care System, 500 Foothill Drive, Salt Lake City, UT 84148
- Saiju Pyarajan Ph.D., Director, Data and Computational Sciences
  VA Boston Healthcare System, 150 S. Huntington Avenue, Boston, MA 02130
- Robert Ringer, Pharm.D., Director, VA Albuquerque Central Biorepository
  New Mexico VA Health Care System, 1501 San Pedro Drive SE, Albuquerque, NM 87108
- Luis E. Selva, Ph.D., Director, MVP Biorepository Coordination
  VA Boston Healthcare System, 150 S. Huntington Avenue, Boston, MA 02130
- Shahpoor (Alex) Shayan, M.S., Director, MVP PRE Informatics
  VA Boston Healthcare System, 150 S. Huntington Avenue, Boston, MA 02130
- Brady Stephens, M.S., Principal Investigator, MVP Information Center
  Canandaigua VA Medical Center, 400 Fort Hill Avenue, Canandaigua, NY 14424
- Stacey B. Whitbourne, Ph.D., Director, MVP Cohort Development and Management
  VA Boston Healthcare System, 150 S. Huntington Avenue, Boston, MA 02130
